# SARS-CoV-2 third vaccine immune response in MS patients treated with ocrelizumab

**DOI:** 10.1101/2022.01.26.22269876

**Authors:** Livnat Brill, Catarina Raposo, Ariel Rechtman, Omri Zveik, Netta Levin, Esther Oiknine-Djian, Dana G. Wolf, Adi Vaknin-Dembinsky

## Abstract

The introduction of a third dose vaccination along with new variants of concerns raises questions regarding serology and T-cell responses in patients with MS (pwMS) treated with B-cell depletion who develop attenuated humoral response to vaccines. The aim of this study is to longitudinally evaluate humoral and cellular response to SARS-CoV-2 mRNA vaccine in ocrelizumab-treated pwMS before and following third vaccine dose. Following the third vaccine dose, patients who are low or non-responders following initial vaccination did not increase antibody titers. In HCs and ocrelizumab-treated pwMS, cellular response decreased 6 months following initial vaccination and increased significantly after the third booster.

## Introduction

Concerns of waning immunity together with the resurgence of the severe acute respiratory syndrome coronavirus 2 (SARS-CoV-2), despite high percent of the population vaccinated with two doses of the mRNA coronavirus disease 2019 (COVID-19) vaccine, led to the recommendation of a third vaccine dose. Patients with MS (pwMS) treated with B-cell depletion treatments such as ocrelizumab are able to generate a robust T-cell response following two doses of the mRNA vaccine ^1-3^. However, an attenuated humoral response was observed ^1, 4, 5^. The cellular response to SARS-CoV-2 is recognizing multiple sites of the receptor-binding domain (RBD) region, thus offering broad reactivity against spike epitopes. Although SARS-CoV-2 variants of concern (VOC) are able to evade infection or vaccination-induced neutralizing antibodies ^6^ a robust T cell response described against VOCs, including the newest Omicron variant was recently described ^7, 8^. Preservation of vaccine-induced T-cell responses may offer some level of protection to individuals with low antibody response. Whether ocrelizumab will have an impact on the longevity of the anti-SARS-CoV-2 response, and the effect of a third dose on the immune response of patients treated with ocrelizumab, are unknown. We assessed longitudinal humoral and T-cell response in HCs and pwMS treated with ocrelizumab pre and post-third BNT162b2 dose. This information can help guide treatment and vaccination recommendations.

## Methods

### Participants and Setting

This single-center study was performed at Hadassah Medical Center, Jerusalem, Israel. Participants were vaccinated between December 2020 and October 2021 and donated blood for antibody and T-cell assessments 2-8 weeks and 6 months following their second dose and 2-8 weeks following the third vaccine dose (BNT162b2, Pfizer/BioNTech). All participants provided written informed consent (975-20 HMO).

### SARS-CoV-2 IgG

Serology response was measured using spike receptor-binding domain (RBD) Architect SARS-CoV-2 IgG II Quant assay (Abbott Diagnostics). Positive and borderline response defined by IgG titer of ≥50 and 50-100 arbitrary units (AU/ml), respectively.

### IFNγ ELISPOT and Cytokine analysis

SARS-CoV-2-specific T-cell response was assessed using T-SPOT^®^ Discovery SARS-CoV-2 (Oxford Immunotec), using freshly isolated peripheral blood mononuclear cells^1^. Results are presented as the number of IFNγ spot-forming cells (SFCs/250,000 cells). Positive response is defined as SFC≥6. IL-2, IL-4, IL-6, IL-10, TNFα and IL-17 levels were measured in supernatants using BD Cytometric Bead Array Human Inflammatory Cytokines Kit.

### Statistical Analysis

Statistical analyses were performed using 1-way ANOVA, student t-test, and Pearson’s correlation coefficient. The results are presented as mean (SD). Two-sided P values are statistically significant at less than 0.05.

## Results

### Participants

A group of 40 HCs (mean [±SD] age 43y±14.8, 24 females) and 33 pwMS treated with ocrelizumab (mean [±SD] age 47.8y±13.5, disease duration: 13.2y±11.4; 23 females, 19 RRMS, 14 PRMS, OCR treatment duration: 28.8±9.6 months and time from last infusion to first and third vaccine 4.9±3.4 and 5±2 months) who received three doses of the BNT162b2 vaccine. The first two doses were given 3 weeks apart, and the third was administered 5-6 months after the second dose.

### SARS-CoV-2 mRNA Vaccine Antibody Response following three doses

Following our publication of attenuated antibody response to SARS-CoV-2 mRNA vaccine in pwMS treated with ocrelizumab ^1^, we evaluated longitudinally the serology response at 6 months post-second dose and 2-8 weeks post-third dose. We observed a significant decline in antibody levels 6 months post-second dose, in both HCs (14352±9453, 1241±885, p<0.0001) and in pwMS treated with ocrelizumab (307.6±604.2, 121.2±329.2, p<0.0001). The third dose increased antibody titers, above 2-8 weeks post-second vaccine in HCs (25851±11302, p<0.0001) and overall, in pwMS treated with ocrelizumab (1413.2± 3742, p=0.9, p=0.007) but in particular in 6/9 initially seropositive patients (814±609.4 vs 6252±6056 p=0.05, 2-8 weeks post-second and post-third dose, respectively). Levels remained positive in 3/9 initially seropositive patients. Three patients with borderline titers and 15 patients with negative antibody response to the first two doses remained negative following the third dose (Fig 1C-D). Titers and response rate post-third dose were significantly lower in pwMS treated with ocrelizumab compared to HCs (p<0.001; 9/27 33.3% vs 30/30 100%, for OCR vs HC, respectively). All patients with increased antibody levels after the third dose were initially vaccinated ≥5 months after the last ocrelizumab infusion. Moreover, patients who were vaccinated with third dose ≥5 months following the last ocrelizumab infusion had a significantly increased likelihood for a positive serologic response (8 of 9 [88.9%] vs 6 of 17 [35.5%]; χ^2^ = 4; *P* = .04) (Figure 1E-F).

**Figure 1:**
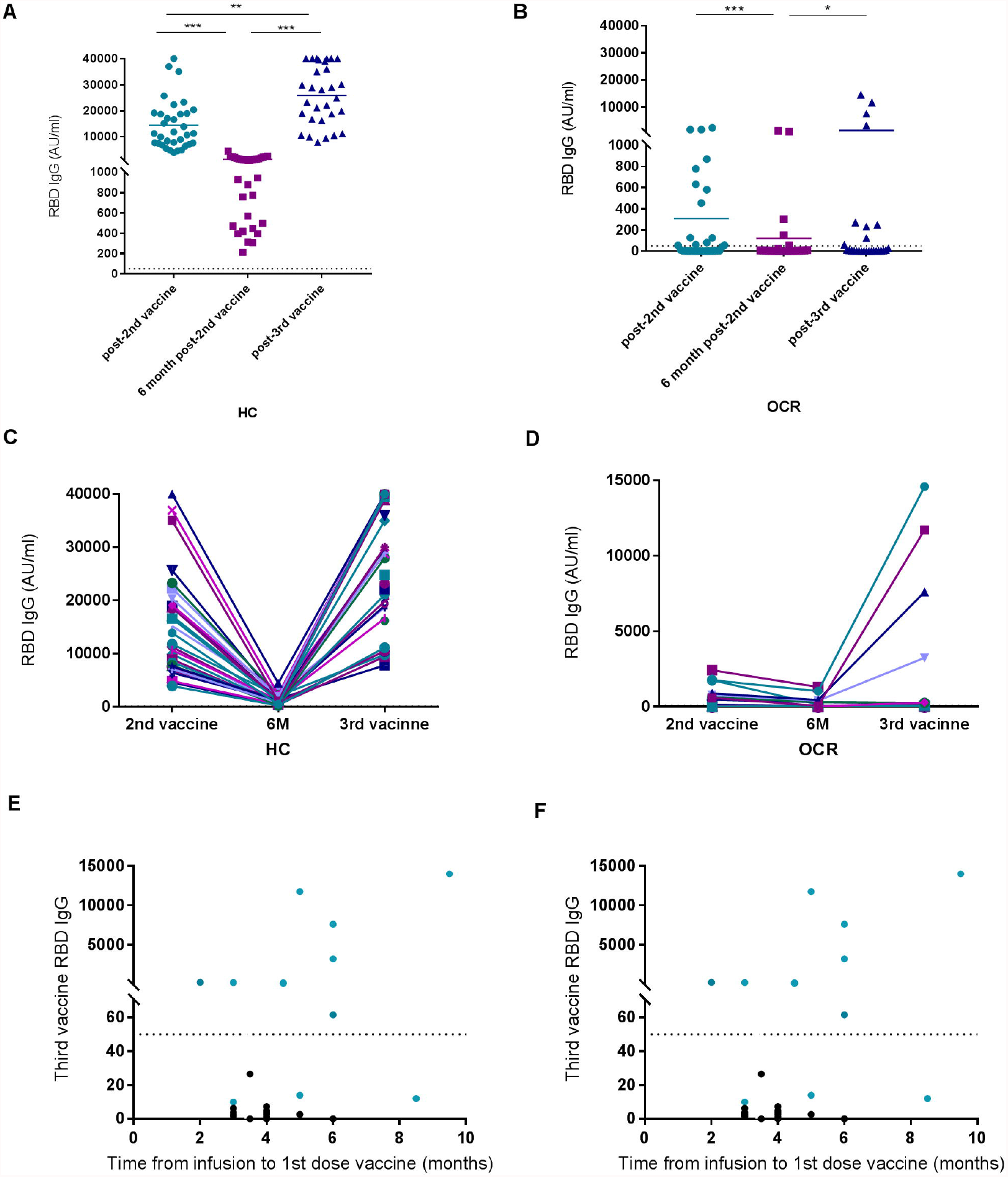
Serology response to the third dose of SARS-CoV-2 mRNA vaccine in patients with MS treated with Ocrelizumab and HCs. (A-B) Longitudinal SARS-CoV-2 RBD-IgG titers in HCs at three time points. 2-4 weeks post-second vaccine dose: n=34, 14352±9453 AU/ml; 6 months post-second vaccine: n=34, 1241±885 AU/ml; and 2-8 weeks post-third vaccine dose: n=30 25851±11302 AU/ml. (B) RBD-Ig titers in 32 HCs, each line represents one participant. (C-D) Longitudinal SARS-CoV-2 RBD-IgG titers in MS patients treated with ocrelizumab at three time points. 2-4 weeks post-second vaccine dose: n=32, 307.6±604.2 AU/ml; 6 months post-second vaccine dose: n=24 121.2±329.2 AU/ml and 2-8 weeks post-third vaccine dose: n=27, 1413±3742 AU/ml. (D) RBD-Ig titers in 32 patients with MS treated with ocrelizumab, each line represents one participant. The dotted line indicates the cut-off for a positive response (≥50 AU/mL). (E-F) Correlation between time from ocrelizumab infusion to vaccination. Colored dots represent seropositivity post second vaccine dose. * p < 0.05; **p < 0.01; ***p < 0.001. Data are presented as mean ± SD.

### Decreased SARS-CoV-2-specific T-cells 6 months following vaccination are boosted with a third dose

T-cell response decreased significantly 6 months following the second vaccine dose (HC: 25.4±10.8, 11±5.4, p=0.004; OCR: 27.4±22.1, 16.3±9.2, p=0.008) and was restored to 2-8 weeks post-second dose levels following the third dose (HC: 26.6±15; OCR: 30.3±21), in both groups. There were no significant differences in response rate and level between HCs and pwMS treated with ocrelizumab post-second and third doses (p=0.5, Figure 2 A-B).

**Figure 2:**
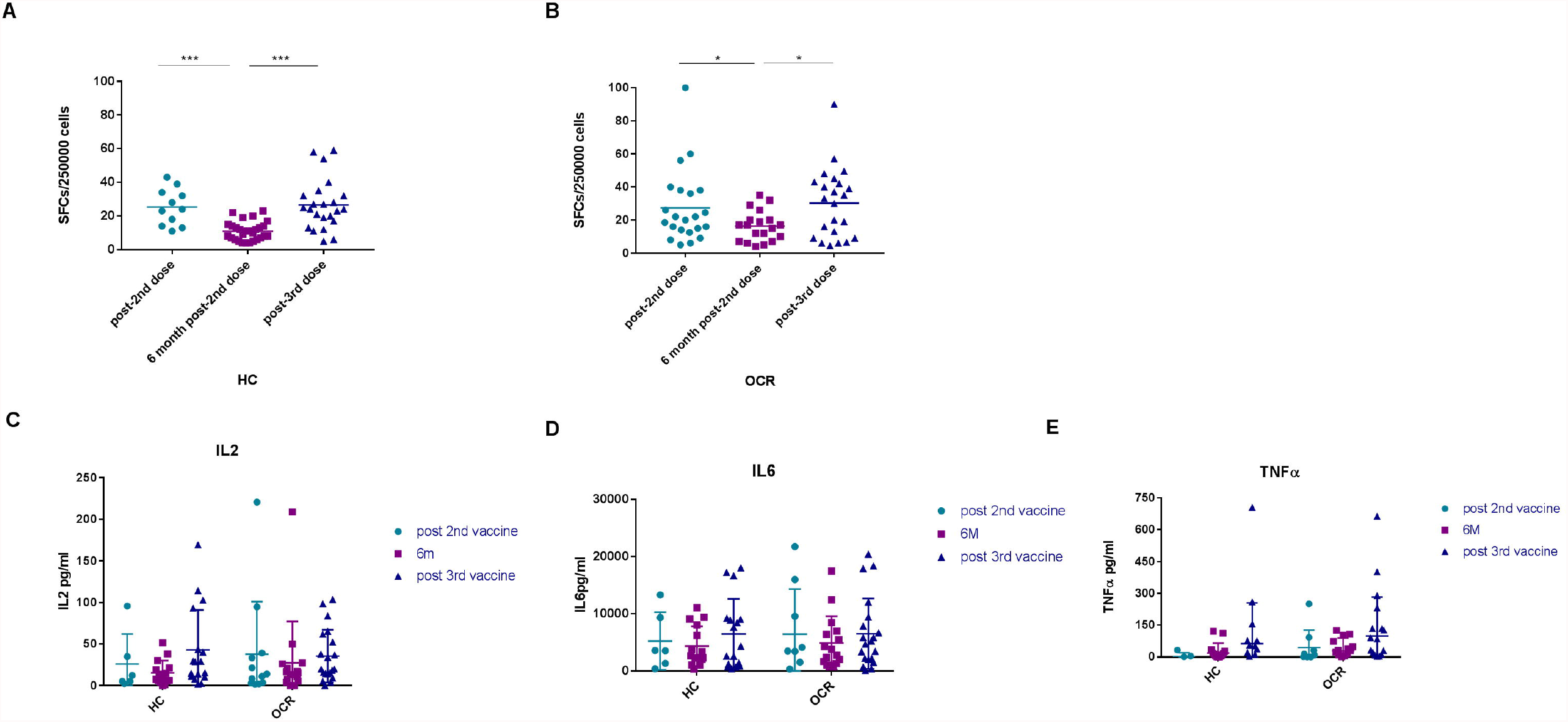
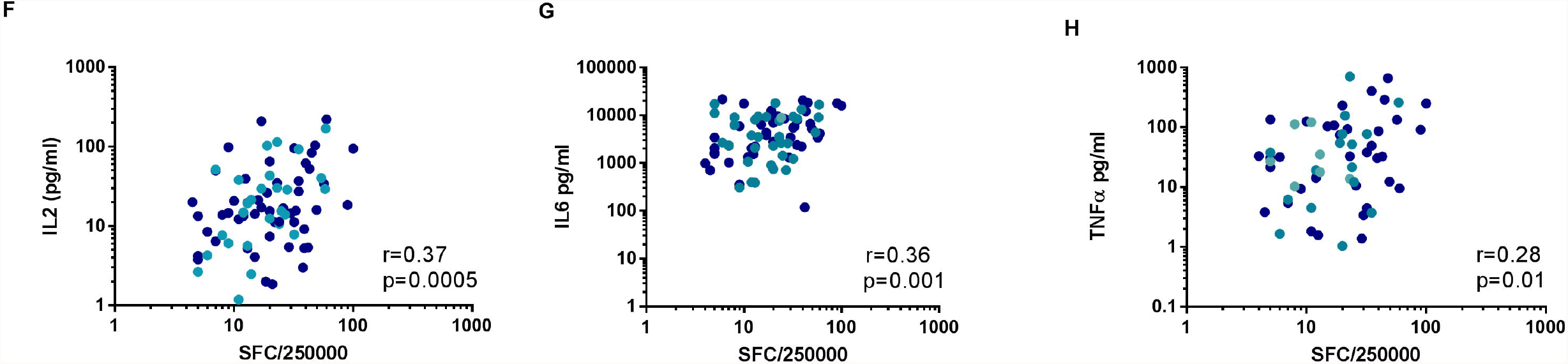
SARS-CoV-2 spike-specific T-cell response following vaccination with three vaccine doses Longitudinal post-vaccination T-cell responses to SARS-CoV-2 peptides. Participants’ PBMCs were stimulated with SARS-CoV-2 spike and nucleocapsid proteins peptides, nil control and phytohemagglutinin control. Response was measured by an interferon-γ ELISpot (T-SPOT, Oxford Immunotec). (A) SARS-CoV-2–specific T-cell response of HCs at three time points. 2-4 weeks post-second vaccine dose: n=11, 25.4±10.8, 6 months post-second vaccine dose: n=29, 11±5.4 and 2-8 weeks post-third vaccine dose: n=23, 26.6±15. (B) SARS-CoV-2–specific T-cell response of pwMS treated with ocrelizumab at three time points. 2-4 weeks post-second vaccine dose: n=22, 27.4±22.1; 6 months post-second vaccine dose: n=19, 16.3±9.2 and 2-8 weeks post-third vaccine dose n=23, 30.3±21. (C-E). Cytokine secretion following stimulation with SARS-CoV-2 spike and nucleocapsid peptides. (cytokine analysis performed after reduction of response to nucleocapsid peptides for each participant). In OCR treated MS patients, IL-2, IL-6 and TNF? levels followed the same pattern as the INF-γ secreting cells, decreased at 6 months after the second vaccine dose and increased with the third vaccine dose (IL2: 37.9±63.3, 28±51.7, 35.6±31.7, p=0.2. IL6: 6426±7878, 4913±4643, 6523±6172; p= 0.34. TNF?: 44.5±83, 29.9±59.8, 99.2±183.8; p=0.6). In HCs, IL-2 and IL-6 levels followed the same pattern of decreased levels 6 month post-second vaccine dose and increased levels following the third vaccine (IL-2: 26±36.2, 15.5±14.6, 42.9±48, p=0.3; IL6: 5247±5028, 4338±3482, 6482±6124, p=0.2; TNF?: 2±18.5, 19.3±46.7, 63.4±192, p= 0.2). (F-H) Correlation between cytokine level and the level of IFNγ-secreting cells. * p < 0.05; **p < 0.01; ***p < 0.001. Data are presented as mean ± SD.

The spike peptides induced secretion of the TH1 cytokine IL-2 and pro-inflammatory cytokines IL-6 and TNFα; with undetected levels of IL-4 and IL-17 and low levels of IL-10. There were no significant differences in cytokine secretion between the two groups (Figure 2C-E). The level of IFN-γ secreting cells significantly correlated with IL-2 in both groups (OCR: r=0.3 p=0.035, HC: r=0.5 p<0.001) and with TNFα and IL-6 levels in pwMS treated with ocrelizumab (r=0.35, p=0.002; r=0.39, 0.01, respectively) (Figure 2F-H).

## Discussion

The need for a third vaccine dose emerged when an increase in breakthrough cases was observed even after vaccination with two doses of mRNA vaccines. Studies revealed waning vaccine immunity over time^9^ and a strong correlation between time of vaccine administration and infection incidence ^10, 11^.

Here, we found that SARS-CoV-2-specific T-cell responses decreased 6 months following vaccination and increased significantly after the third dose, both in HCs and pwMS treated with ocrelizumab. Antibody levels also significantly decreased 6 months after the first two doses in both groups, and significantly increased following the third dose in HCs and in a subset of seropositive pwMS treated with ocrelizumab. Data on the effect of a third vaccine dose on people with negative serology is scarce^12^. In organ transplantation it has been reported that 44% of the seronegative patients before the third dose were seropositive afterwards^13^. In a small cohort of rheumatology patients treated with another B-cell-depletion therapy, rituximab, vaccinated seronegative patients remained seronegative after the third vaccine dose with preserved T-cell response^14^. We found that none of the borderline/seronegative patients developed positive serology response post-third dose. All patients with increased antibody levels after the third dose were vaccinated more than five months after the last ocrelizumab infusion, suggesting that it may be possible to optimize the antibody response to vaccination by timing vaccination with regards to the last infusion dose.

The 6-month decline and the post booster increase in T-cell response was similar in HC and pwMS-treated with ocrelizumab providing initial evidence that ocrelizumab may not affect the longevity of the T-cell response. While T-cells have been suggested to have a protective role, in particular in controlling disease severity^15^ they also seem to be less influenced from variants that escape antibody neutralization^8^. Omicron, the most recent VOC with a high number of mutations in the receptor binding domain, has been shown to have significant evasion of vaccine-induced neutralizing antibody responses^16, 17^. Most of the experimental T-cell epitopes known to be targeted in vaccinated and/or previously infected are unaffected by Omicron mutations^18^. This highlights the importance of T-cell response following vaccination. Evidence from breakthrough infection will help understand the contribution of antibodies and T-cells in mediating protection from severe COVID-19. A recent study suggested that anti-CD20s-treated patients may be at higher risk of COVID-19 infection^19^. Nevertheless, the cases were mostly mild, with most patients not requiring hospitalization, none required ICU admission, and none died.

Our findings show that approximately 6 months after 2 doses of BNT162b2, both HCs and pwMS on OCR had decreased levels of SARS-CoV-2 antibodies and T cells and underscore the benefit of a third dose boosting the immune response (antibodies and/or T cells). Follow-up studies will help address whether a third dose provides enhanced long-term protection, effect on the quality of the immune response and waning rate compared to two doses, in the general population and in pwMS.

In conclusion, a third vaccine restored T-cell levels in both pwMS treated with ocrelizumab and HCs. For seronegative patients, a third dose did not increase antibody titers. A small fraction of ocrelizumab-treated patients had boosted antibody titers. Additional studies are needed to optimize vaccine response in patients treated with anti-CD20s. Our findings support the administration of a third vaccine dose in pwMS taking ocrelizumab.

## Data Availability

All data produced in the present study are available upon reasonable request to the authors

## Author Contributions

Drs Vaknin-Dembinsky and Brill had full access to all of the data in the study and take responsibility for the integrity of the data and the accuracy of the data analysis.

Concept and design: Brill, Raposo, Vaknin-Dembinsky.

Acquisition, analysis, or interpretation of data: Brill, Rechtman, Zveik, Oiknine-Djian, Wolf, Levin, Raposo, Vaknin-Dembinsky.

*Drafting of the manuscript:* Brill, Raposo, Vaknin-Dembinsky.

*Critical revision of the manuscript for important intellectual content:* Brill, Raposo, Vaknin-Dembinsky.

*Statistical analysis:* Brill.

*Administrative, technical, or material support:* Brill, Raposo, Vaknin-Dembinsky.

*Supervision:* Wolf, Levin, Vaknin-Dembinsky.

